# Effect of transcranial direct current stimulation on post-stroke fatigue

**DOI:** 10.1101/2020.11.18.20227272

**Authors:** William De Doncker, Sasha Ondobaka, Annapoorna Kuppuswamy

## Abstract

**Background:** Fatigue is one of the most commonly reported symptoms post-stroke, which has a severe impact on quality of life. Post-stroke fatigue is associated with reduced motor cortical excitability, specifically of the affected hemisphere.

**Objective:** The aim of this exploratory study was to assess whether fatigue symptoms can be reduced by increasing cortical excitability using anodal transcranial direct current stimulation (tDCS).

**Methods:** In this sham-controlled, double-blind intervention study, tDCS was applied bilaterally over the primary motor cortex in a single session in thirty stroke survivors with high severity of fatigue. A questionnaire-based measure of trait fatigue (primary outcome) was obtained before, after a week and a month post stimulation. Secondary outcome measures of state fatigue, motor cortex neurophysiology and perceived effort were also assessed pre, immediately post, a week and a month post stimulation.

**Results:** Anodal tDCS significantly improved fatigue symptoms a week after real stimulation when compared to sham stimulation. There was also a significant change in motor cortex neurophysiology of the affected hemisphere and perceived effort, a week after stimulation. The degree of improvement in fatigue was associated with baseline anxiety levels.

**Conclusion:** A single session of anodal tDCS improves fatigue symptoms with the effect lasting up to a week post stimulation. tDCS may therefore be a useful tool for managing fatigue symptoms post-stroke.

## Introduction

Debilitating fatigue that persists for months and sometimes years after stroke is relatively common in stroke survivors with prevalence as high as 70%[1,2]. Post-stroke fatigue (PSF) has been identified as the top unmet need among stroke survivors living in the community and is a top priority for further research[3–5].

Despite the high prevalence of fatigue, the pathophysiology of chronic fatigue is poorly understood with little evidence-based therapy to alleviate fatigue[6]. Our recent work aimed at understanding the underlying neurophysiology of PSF provided a potential target for modulation that may reduce the symptoms of PSF. We showed using transcranial magnetic stimulation (TMS), that cortical excitability at rest, specifically the primary motor cortex (M1) of the affected hemisphere, is reduced in those who report high levels of PSF[7]. Motor cortex excitability as measured by TMS is normally associated with motor function of the targeted muscle[8]. However, TMS measures of motor cortex excitability have also been associated with non-motor functions such as perception and attention[9,10]. Given the homogeneity of motor function in the investigated stroke cohort, we argued that reduced motor cortex excitability was a reflection of altered perceptual processing in relation to muscle contraction i.e. altered effort perception.

Perceived effort (PE) is heavily influenced by both expectations and feedback, efferent and afferent input. The active inference framework of sensorimotor control provides a simple framework that integrates both efferent and afferent input to explain motor control[11–19]. Within this framework, expectations set the gain for afferent input and PE is a psychophysical output of the gain function[20]. We recently showed that those with greater PSF did indeed show greater PE[21]. Increased PE may be driven by altered gain, as measured by motor cortex excitability[20]. We hypothesised that altering the gain i.e. increasing motor cortex excitability, will reduce PE and subsequently reduce fatigue symptoms.

Transcranial direct current stimulation (tDCS) is a non-invasive brain stimulation technique that increases or decreases cortical excitability when applied to the M1, depending on the montage and stimulation parameters used[22,23]. tDCS has promising potential therapeutic applications due to its ease of use, low cost and lack of physiological and behavioural side effects[24,25]. A single session of tDCS for a few minutes can result in after effects that last for more than an hour, with within session repeated tDCS resulting in more pronounced and longer lasting effects[26,27]. tDCS modulates cortical excitability in stroke survivors and has been widely used in the treatment of various neurological and psychiatric disorders, including the treatment of multiple sclerosis fatigue[28–37]. tDCS has also been used to enhance sport performance and has been shown to reduce rate of perceived exertion in healthy individuals[38].

The primary aim of this exploratory study was to use anodal tDCS over the M1 to reduce PSF. The secondary aim was to investigate the potential mechanisms that underlie the hypothesised effect on PSF.

## Methods

### Study design

This was a double-blind, sham-controlled study with a single session of bilateral anodal tDCS chosen as the method of intervention. Patients visited the laboratory on three separate occasions, with tDCS applied only on the first visit. The second visit took place one week later with the third visit taking place one month after visit two (figure 1). The primary outcome measure was a change in trait fatigue. Secondary outcome measures included state fatigue, explicit and implicit measures of PE and motor cortex physiology measures of resting motor thresholds (RMT) and slope of recruitment curves (IO_Slope_) of the affected and un-affected hemisphere assessed using TMS. The primary outcome measure was recorded at three distinct time points (pre stimulation, week and month post stimulation). All other outcome measures were recorded at four distinct time points (pre stimulation, immediately post stimulation, week and month post stimulation).

**Figure 1.**
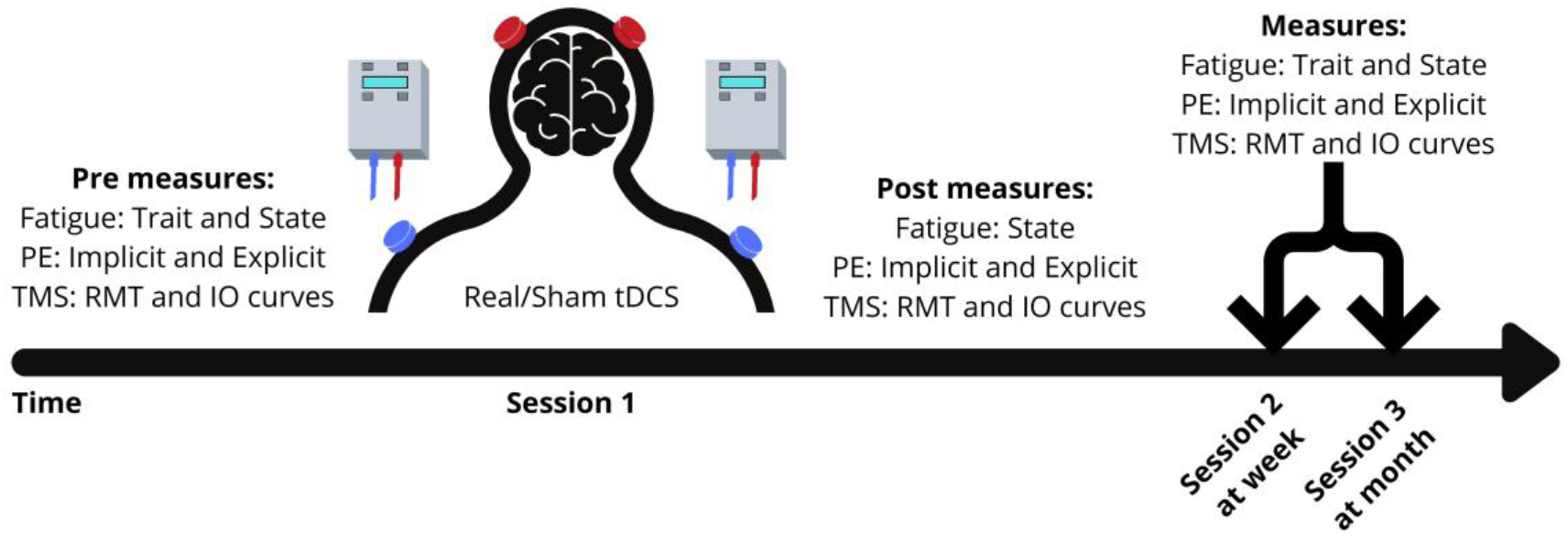
Study Design indicating the sequence in which procedures where done for each of the sessions at the 4 different time points (pre tDCS, immediately post-tDCS, week post-tDCS and month post-tDCS).

### Subjects

The study was approved by London Bromley Research Ethics Committee (16/LO/0714). Stroke survivors were recruited via Clinical Research Network at University College NHS Trust Hospital, a departmental Stroke Database and from the community.

Inclusion criteria: Date of stroke > 3 months, first time stroke, age ≥ 18 years; Fatigue Severity Scale (FSS-7) ≥ 4. A score of ≥ 4 on FSS-7 indicates the presence of clinically significant fatigue[39]. Exclusion criteria: use of centrally-acting medication which may affect the level of fatigue, depression, and anxiety; depression scores ≥ 11 (Hospital Anxiety and Depression Scale - HADS); grip strength and manual dexterity (nine-hole peg test) ≤ 60% of unaffected hand.

The minimal clinically important difference on the FSS-7 is 0.45, with differences greater than 0.45 predicting a significant effect on quality of life[40,41]. To detect the minimal clinically important difference in fatigue with 80% power (0.80) and a significance level alpha of 0.05, a sample size of 11 subjects per group is needed. Twice the number of patients were allocated to the real stimulation group than necessary, as previous studies using tDCS in multiple sclerosis fatigue and healthy individuals showed that approximately 50% of patients respond to tDCS[31,42]. Thirty-three patients were recruited into the study and were randomly allocated to the real (n = 22) or sham (n = 11) stimulation groups (figure 2). All patients gave written informed consent in accordance with the Declaration of Helsinki. Patient demographics for both groups are found in table 1.

**Table 1.**
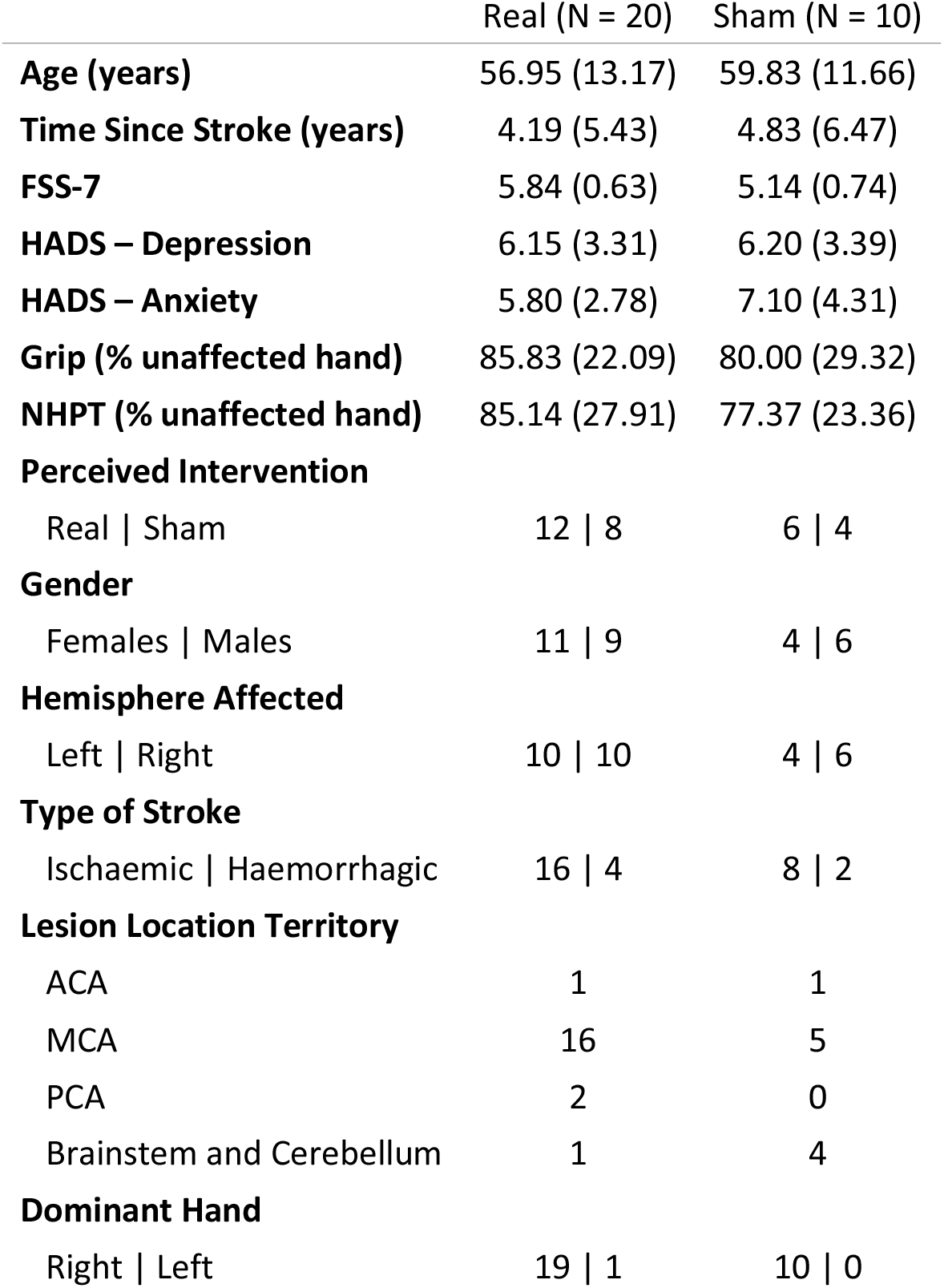
Patient demographics and clinical data for the real and sham stimulation groups. FSS-7 = Fatigue severity Scale-7; HADS = Hospital Anxiety and Depression Scale; NHPT = Nine-hole peg test; ACA = Anterior Cerebral Artery; MCA = Middle Cerebral Artery; PCA = Posterior Cerebral Artery.

|  | Real (N = 20) | Sham (N = 10) |
| --- | --- | --- |
| **Age (years)** | 56.95 (13.17) | 59.83 (11.66) |
| **Time Since Stroke (years)** | 4.19 (5.43) | 4.83 (6.47) |
| **FSS-7** | 5.84 (0.63) | 5.14 (0.74) |
| **HADS – Depression** | 6.15 (3.31) | 6.20 (3.39) |
| **HADS – Anxiety** | 5.80 (2.78) | 7.10 (4.31) |
| **Grip (% unaffected hand)** | 85.83 (22.09) | 80.00 (29.32) |
| **NHPT (% unaffected hand)** | 85.14 (27.91) | 77.37 (23.36) |
| **Perceived Intervention** |  |  |
| Real \| Sham | 12 \| 8 | 6 \| 4 |
| **Gender** |  |  |
| Females \| Males | 11 \| 9 | 4 \| 6 |
| **Hemisphere Affected** |  |  |
| Left \| Right | 10 \| 10 | 4 \| 6 |
| **Type of Stroke** |  |  |
| Ischaemic \| Haemorrhagic | 16 \| 4 | 8 \| 2 |
| **Lesion Location Territory** |  |  |
| ACA | 1 | 1 |
| MCA | 16 | 5 |
| PCA | 2 | 0 |
| Brainstem and Cerebellum | 1 | 4 |
| **Dominant Hand** |  |  |
| Right \| Left | 19 \| 1 | 10 \| 0 |

**Figure 2.**
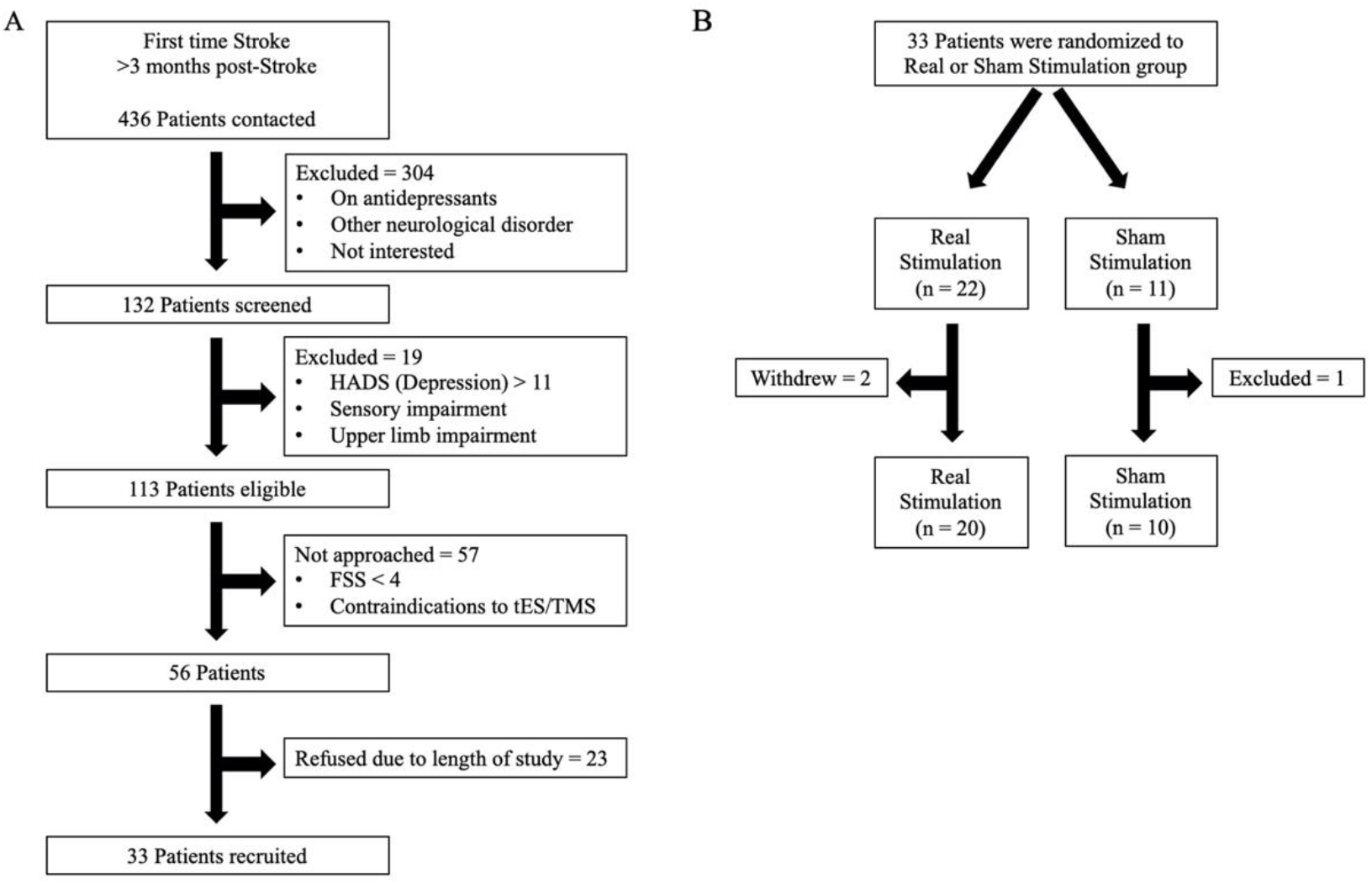
Study recruitment and randomisation (HADS = Hospital Anxiety and Depression Scale; FSS = Fatigue Severity Scale; tES = transcranial Electrical Stimulation; TMS = Transcranial Magnetic Stimulation).

### Questionnaires

Trait and state measures of fatigue were captured during the study. Trait fatigue represents the experience and impact of fatigue on day to day living for a pre-determined time period leading up to the day of testing, whereas state fatigue characterizes fatigue at a given moment in time. Trait fatigue was quantified using the FSS-7, a seven-item questionnaire asking for ratings of fatigue ranging from one to seven (strongly disagree to strongly agree) over the preceding week from the day of administration[43]. An average score of one indicates no fatigue while an average score of seven indicates very severe fatigue. State fatigue was quantified using a visual analogue scale (VAS) ranging from zero to ten (Not at all tired to extremely tired). Patients also completed the HADS, a 14-item questionnaire with a depression and anxiety subscale, prior to the stimulation. A score of 0 to 7 for either subscale could be regarded as being in the normal range, with a score of 11 or higher indicating probable presence of the mood disorder[44].

### Stimulation

tDCS was applied using two battery-driven stimulators (DC-Stimulator Plus, NeuroConn, Germany) while patients were awake and at rest. Four 35 cm^2^ rubber electrodes coated with conductive paste were secured with self-adhesive bandages. The anode of each stimulator was placed over the left and right M1 (C3 and C4 according to the 10-20 EEG system), while the cathode of each stimulator was placed over the ipsilateral left and right shoulders. This tDCS montage has previously been shown to reduce perception of effort and increase corticospinal excitability in healthy individuals[38]. Real tDCS involved two 20-minute sessions of stimulation at 2 mA separated by a 10-minute break in between. The current was ramped up for 30 seconds until reaching 2 mA and ramped down for 30 seconds at the end of the stimulation period. Stimulation intensity and duration complied with current safety recommendations[45]. For sham stimulation, the current was ramped down immediately after ramping up, providing effective blinding[46]. The patient and researchers were blind to the applied stimulation (real or sham). At the end of stimulation, patients were explicitly asked whether they thought they received real or sham stimulation.

### Perceived effort

PE was measured in an isometric hand grip task with a hand-held dynamometer (Biometrics Ltd, Newport, UK) performed using the dominant hand as in De Doncker et al[21]. Measures of implicit and explicit PE were obtained using a line length estimation task and a VAS respectively.

### Surface electromyogram and TMS

Electromyogram (EMG) recordings were obtained from the first dorsal interosseous (FDI) muscle using surface electrodes (1041PTS Neonatal Electrode, Kendell) in a belly-tendon montage with the ground positioned over the flexor retinaculum of the hand. The signal was amplified with a gain of 1000 (D360, Digitmer, Welwyn Garden City, UK), bandpass filtered (100-1000 Hz), digitized at 10kHz (Power1401, CED, Cambridge, UK) and recorded with Signal version 6.04 software (CED, Cambridge, UK).

TMS (figure-of-eight coil with wing diameter, 70mm; Magstim 2002, Magstim, Whitland, UK) was used to stimulate the hand area of the M1. The coil was held tangentially on the scalp at 45^°^ to the mid-sagittal plane to induce a posterior-anterior current across the central sulcus. The subjects were instructed to stay relaxed with their eyes open and their legs uncrossed. The motor ‘hotspot’ of the FDI muscle was determined as described previously[7]. RMT was defined as the lowest intensity required to evoke a motor evoked potential (MEP) at the hotspot of at least 50μV in a minimum of 5 of 10 consecutive trials while subjects were at rest. IO curves were acquired at rest at the hotspot using TMS intensities set at 100, 110, 120, 130, 140 and 150% of RMT. Six pulses at each of the 6 intensities were delivered in a randomized order with an inter-trial interval of 4 seconds. This procedure was repeated for both the affected (RMT-A, IO_Slope_-A) and unaffected (RMT-U, IO_Slope_-U) hemispheres.

### Analysis of questionnaires

The FSS-7 score was calculated by averaging all items for each of the three time points.

The total score was taken for the anxiety and depression subscales of HADS, HADS-Anxiety and HADS-Depression respectively, and were considered as independent measures.

### TMS analysis

The data were analysed using custom-written routines in Matlab (2018a, Mathworks). Peak-to-peak MEP amplitudes for each condition were estimated from the EMG recordings. All trials were visually inspected and approximately 7% of trials with pre-contraction and size ≤ 0.025 mV were excluded across all participants. The median MEP amplitude for each participant at each TMS intensity (100-150% RMT) was calculated. IO curve measurements for each session were estimated by applying linear fits to the resultant values (mean R^2^ = 0.86). For each hemisphere (affected and un-affected hemisphere) the gradient of the linear fit was calculated for each of the four time points.

### PE analysis

To obtain a measure of explicit PE, VAS scores were averaged across all trials in each force level in each individual. As there was no difference across force level, the average VAS score across all force levels was used as an explicit measure of PE for each of the four time points. The sum of the number of lines reported as long for each individual in each force level was calculated. As there was no difference across force level, the average number of lines across all force levels was used as an implicit measure of PE for each of the four time points.

### Statistical analysis

All statistical analysis was performed using R (RStudio Version 1.2.5033). Assumptions of a normal distribution of the primary and all secondary outcome variables was assessed using the Shapiro-Wilk test. All data were non-normally distributed (p < 0.05). To test for changes over time, a non-parametric Friedman test was performed for the primary outcome variable (trait fatigue) and all secondary outcome variables (state fatigue, RMT-A, RMT-U, IO_Slope_-A, IO_Slope_-U, PE-implicit and PE-explicit), separately for the sham and real intervention groups as in Saiote et al[31]. When significant results were found, pairwise comparison between baseline and each post-measurement day were performed using Wilcoxon signed-rank test. To analyse the effect of real stimulation versus sham stimulation across both primary and secondary outcome measures, the changes in scores were calculated by normalising each day to baseline (pre stimulation), and then compared within each day using a Wilcoxon signed-rank test. Adjustment for multiple comparisons was performed using Bonferroni correction.

A spearman correlation was used to examine the association between baseline trait fatigue scores and the change in trait fatigue a week after stimulation in both the sham and real stimulation groups. To identify the potential mechanisms that drive the change in trait fatigue in the real stimulation group a multiple linear regression was used with demographic data and secondary outcome variables that were significantly different between the real and sham stimulation groups used as predictors. Goodness of fit was assessed using the BIC (lower BIC indicates a better fitting model) to identify the combination of variables that best predicted the outcome variable, the change in trait fatigue. Assumptions of normality and homoscedasticity of the residuals for each model were assessed visually using quantile-quantile normal plots and fitted-versus residual-value plots.

## Results

Two patients from the real stimulation group decided to withdraw from the study during visit 1 as they found the stimulation uncomfortable and one patient from the sham stimulation group was excluded because they started taking antidepressants after session 1. No serious adverse events were reported. Thirty patients were included in the final analysis, with twenty in the real stimulation group and ten in the sham stimulation group. Only 60% of patients in both the real and sham stimulation groups correctly predicted the type of stimulation they received, suggesting that our blinding was effective.

### Trait and State Fatigue

The Friedman test showed a significant effect of time on trait fatigue in the real stimulation group (χ^2^(2)=15.5; p<0.001) but not in the sham stimulation group (χ^2^(2)=0.154; p=0.926). Post-hoc analysis with Wilcoxon signed-rank tests showed that FSS-7 scores decreased compared to baseline both at the week (V=198, Z=0.777, p=0.002) and month (V=142, Z=0.535, p=0.047) time point in the real stimulation group. FSS-7 at the week time point was also lower than at the month time point (V=21, Z=0.639, p=0.016). Post-hoc Wilcoxon signed-rank test of the normalised FSS-7 scores revealed a significant difference between the sham and real stimulation group at the week time point (W=52.5, Z=0.382, p=0.0386) but not at the month time point (W=65.5, Z=0.277, p=0.134), seen in figure 3A.

**Figure 3.**
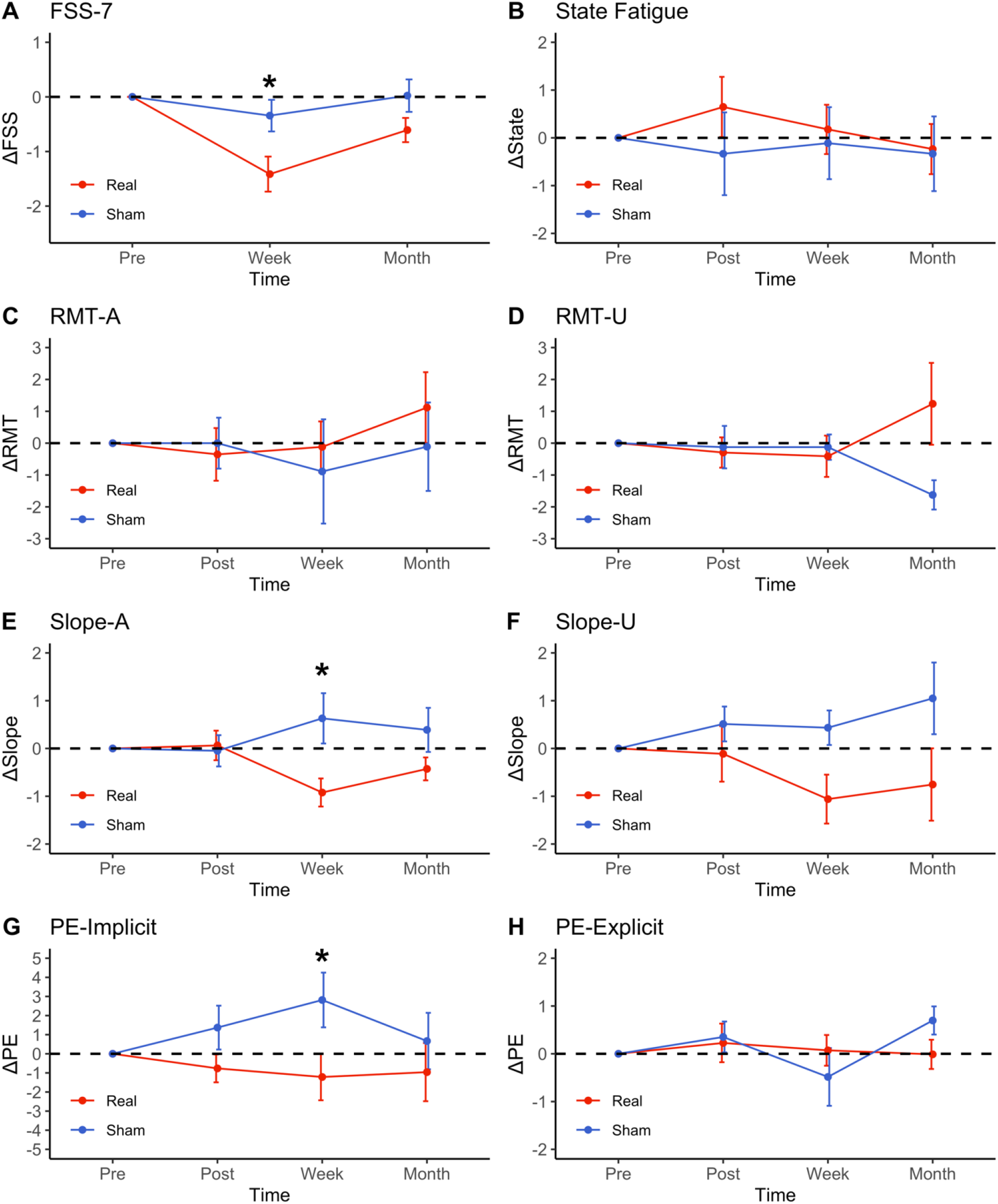
Changes in trait fatigue (A), state fatigue (B), resting motor threshold of the affected and unaffected hemisphere (C-D), slope of the recruitment curve of the affected and unaffected hemisphere (E-F), implicit perceived effort (G) and explicit perceived effort (H) compared to baseline (pre stimulation time point) across the different time points for both the real (red) and sham (blue) stimulation group. Error bars represent standard error of the means. Significance levels are indicated by * (p < 0.05). FSS-7 = Fatigue Severity Scale-7; RMT-A = Resting Motor Threshold of Affected Hemisphere; RMT-U = Resting Motor Threshold of Un-affected Hemisphere; IO_Slope_-A = recruitment curve slope of Affected Hemisphere; IO_Slope_-U = recruitment curve slope of Un-affected Hemisphere; PE = Perceived Effort.

The Friedman test showed no significant effect of time on state fatigue in the real (χ^2^(3)=0.97; p=0.809) or sham (χ^2^(3)=3; p=0.392) stimulation group. Post-hoc Wilcoxon singed-rank test for the normalised state fatigue scores revealed no significant difference between the sham and real stimulation group at any time point (figure 3B).

### Neurophysiology

The Friedman test showed a significant effect of time on IO_Slope_-A in the real stimulation group (χ^2^(3)=11.2, p=0.0106) but not in the sham stimulation group (χ^2^(3)=0.75, p=0.861). Post-hoc analysis with Wilcoxon signed-rank tests showed that IO_Slope_-A decreased compared to baseline at the week (V=96, Z=0.730, p=0.024) but not at the immediate post and month time point in the real stimulation group. Post-hoc Wilcoxon signed-rank test of the normalised IO_Slope_-A scores revealed a significant difference between the sham and real stimulation group at the week time point (W=23, Z=0.480, p=0.024) but not at the immediate post and month time point. The Friedman test showed no significant effect of time on RMT-A in the real (χ^2^(3)=2.62, p=0.454) or sham (χ^2^(3)=2.06, p=0.560) stimulation groups. Post-hoc Wilcoxon singed-rank test for the normalised RMT-A revealed no significant difference between the sham and real stimulation group at any time point.

The Friedman test showed no significant effect of time on RMT-U or IO_Slope_-U in the real (χ^2^(3)2.71, p=0.438; χ^2^(3)=3.38, p=0.337) or sham (χ^2^(3)=5.62, p=0.132; χ^2^(3)=2.2, p=0.552) stimulation groups. Post-hoc Wilcoxon singed-rank test for the normalised RMT-U and IO_Slope_-U scores revealed no significant difference between the sham and real stimulation group at any time point (figure 3C-3F).

### Perceived Effort

The Friedman test showed no significant effect of time on PE-implicit or PE-explicit in the real (χ^2^(3)=5.60, p=0.905; χ^2^(3)=0.2, p=0.978) or sham (χ^2^(3)=4.24, p=0.237; χ^2^(3)=6.73, p=0.0809) stimulation groups. Post-hoc Wilcoxon singed-rank test for the normalised PE-explicit scores revealed no significant difference between the sham and real stimulation group at any time point. Post-hoc Wilcoxon singed-rank test for the normalised PE-implicit scores revealed a significant difference between the sham and real stimulation group at the week time point (W=39.5, Z=0.391, p=0.0491) but not at the immediate post and month time point (figure 3G-3H).

### Change in trait fatigue

There was no association between baseline FSS-7 scores and the change in FSS-7 score at the week time point in the real (r_s_= -0.2, p=0.39) and sham (r_s_=0.2, p=0.57) stimulation group (figure 4A). The multiple linear regression model with normalised IO_Slope_-A at the week time point and baseline HADS-Anxiety scores as predictors was the best fitting model (ΔFSSweek= -2.775+0.436ΔIO_Slope_-A+0.285HADS-Anxiety). A significant regression equation was found (F(2,13)=5.345, p=0.020), with an adjusted R^2^ of 0.37. Baseline HADS-Anxiety significantly explained the change in FSS-7 scores (t=2.925, p=0.013), whereas normalised IO_Slope_-A at the week time point (t=1.760, p=0.104) was not a significant predictor of the change in FSS-7 scores (figure 4B). Beta coefficients of the predictors used in the model together with their associated 95% confidence intervals and p-values are found in table 2.

**Table 2.**
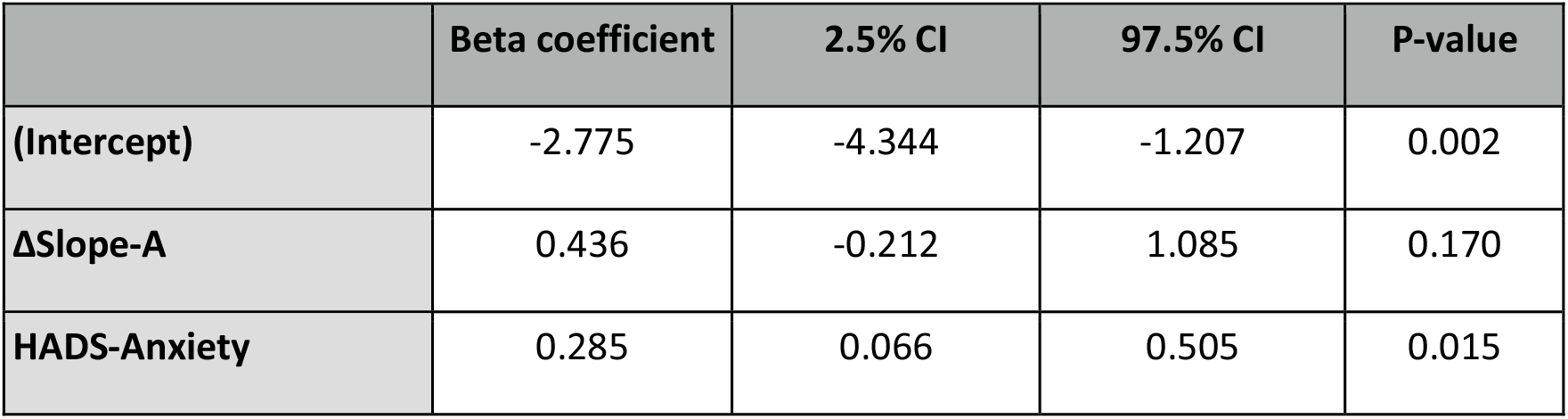
Multiple linear regression results with change in FSS-7 at the week time point as the outcome variable and change in IO_Slope_-A and HADS-Anxiety as predictors. Table includes beta coefficients, 95% confidence intervals and the associated p-values for each predictor. FSS-7 = Fatigue Severity Scale-7; IO_Slope_-A = recruitment curve slope of the affected hemisphere; HADS = Hospital Anxiety and Depression Scale; CI = confidence interval.

|  | Beta coefficient | 2.5% CI | 97.5% CI | P-value |
| --- | --- | --- | --- | --- |
| **(Intercept)** | -2.775 | -4.344 | -1.207 | 0.002 |
| **ΔSlope-A** | 0.436 | -0.212 | 1.085 | 0.170 |
| **HADS-Anxiety** | 0.285 | 0.066 | 0.505 | 0.015 |

**Figure 4.**
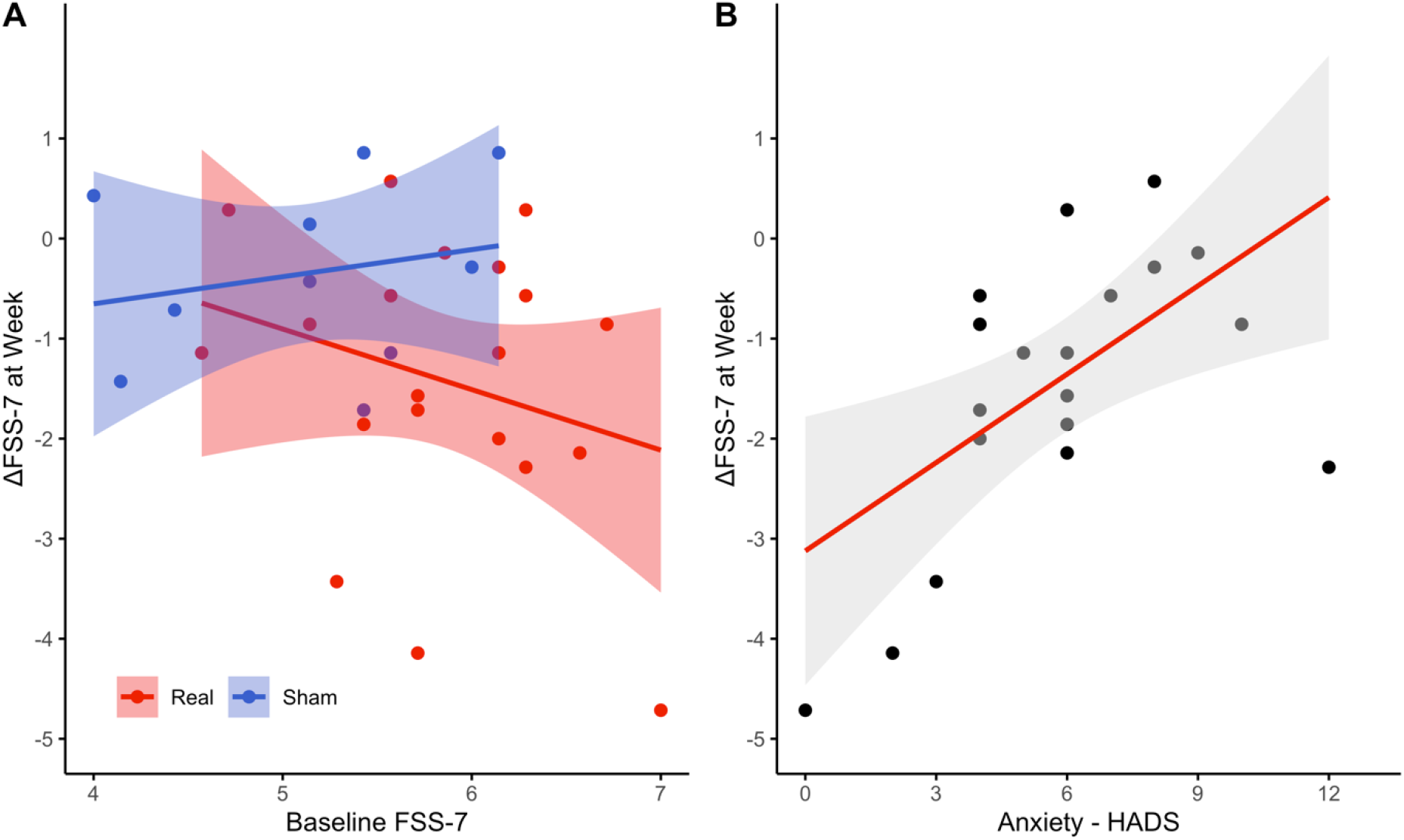
Correlation between baseline FSS-7 and the change in FSS-7 at the week time point for the real (red) and sham (blue) stimulation groups with the 95% confidence interval (A). The association between the baseline HADS-Anxiety levels and the change in FSS-7 at the week time point for the real stimulation group with its associated 95% confidence interval (B). FSS-7 = Fatigue Severity Scale-7; HADS = Hospital Anxiety and Depression Scale.

## Discussion

In this study, we aimed to improve fatigue symptoms in minimally impaired, non-depressed stroke survivors using bilateral anodal tDCS over the M1. We show a significant reduction in trait fatigue a week following anodal tDCS. There were also significant differences between the real and sham stimulation groups in IO_Slope_ of the affected hemisphere and implicit measures of PE a week after stimulation. Those with the greatest drop in trait fatigue a week after anodal tDCS also had the lowest anxiety scores prior to stimulation.

### tDCS and fatigue

Anodal tDCS increases cortical excitability [28] and reduce perception of effort in healthy participants during an endurance performance task[38]. The effects of tDCS are not specific to the targeted brain regions but spread to distinct cortical and subcortical structures[47]. A single session of anodal tDCS was expected to increase motor cortex excitability temporarily resulting in reduction of PE by modulating excitability and connectivity of regions upstream of M1[12,13,48]. By resetting PE in a physical task, the carry over effects on behaviour would accumulate eventually reducing fatigue levels. This reduction in fatigue levels would be evident by changes in trait fatigue but not necessarily state fatigue. The current results partly support this theory by showing a reduction in trait fatigue a week post-tDCS. Fatigue has been identified as one of the top unmet needs for chronic stroke survivors within the community and interferes most with their activities of daily living[3,4]. Currently there are no effective interventions for PSF[49,50]. The proposed intervention, if confirmed to be effective in larger and a more heterogeneous cohort of patients, will represent a simple, low-cost and risk-free procedure for reducing fatigue symptoms[24,30].

### Neurophysiology

RMT is a measure of motor corticospinal excitability and anodal tDCS reduces RMT[28]. The lack of effect of tDCS on RMT in this study could have been driven by our split stimulation paradigm of two 20-minute periods invoking homeostatic metaplasticity[51], with the second 20-minute stimulation reversing the effects of the first 20 minutes. The slope of IO curves following real tDCS reduced significantly in the affected hemisphere when compared to sham tDCS. IO curves are thought to measure the excitability of pathways upstream of motor cortex, therefore a measure of inputs to the motor cortex[52,53]. The slope of IO curves represents the gain of descending corticospinal tract, possibly driven from higher motor areas[54,55]. However, a reduction and not an increase in slope is puzzling. Steeper IO represent greater recruitment of higher order motor areas such as the supplementary motor area and pre motor cortex[53]. As there are no normative values for IO curves, it is hard to speculate if tDCS induced shallowing of IO curves is a reflection of homeostatic normalisation of inputs to the motor cortex, but offers one possible explanation of the IO results.

### Perceived effort

PE has mostly been tested in post-exercise paradigms where measures such as VAS and Borg Scales have been validated against physiological measures of exertion such as heart rate and maximal aerobic capacity[56,57]. They conclude PE is less subject to bias in some populations[58,59]. However, in the current study PE is measured in a non-exercise paradigm and in a disease population with a condition that is highly stigmatised and under recognised[60,61]. Hence, we expected to see a response bias in PE. Therefore, in addition to an explicit measure of PE, we introduce a novel implicit measure of PE based on line length perception. This measure takes advantage of the susceptibility of visual perception to physical effort where high effort unfavourably biases distance estimation[62]. On similar lines, a line length estimation task was developed and shown to be biased by prior exertion, which we use as a measure of implicit PE[21,63]. We showed that trait fatigue is explained by implicit PE but not explicit PE in a physical task[21]. In the current study we show a significant difference between real and sham stimulation in implicit PE but not in explicit PE a week after anodal tDCS. It could be that implicit, perceived effort is the first to respond to tDCS and if the effects were maintained, would result in a reduction in explicit PE. A second possible explanation is that reducing implicit PE that is sufficient to alleviate fatigue. This difference appears to be primarily driven by an increase in implicit PE in the sham stimulation group rather than a decrease in implicit PE in the real stimulation group. The test-retest reproducibility of the paradigm used to measure implicit PE has not been examined in the absence of tDCS, and could potentially shed light on the current finding.

### Mechanism driving the change in trait fatigue

One of the aims of this work was to identify the potential mechanisms that underlie the reduction in trait fatigue following anodal tDCS. The expectation was that a change immediately post stimulation in M1 neurophysiology and PE will result in reduction in trait fatigue a week later. However, no change immediately post stimulation, but only a week later in these measures, makes it harder to interpret the neurophysiology and perception results. The results of the multiple linear regression model suggest that measures of M1 neurophysiology, specifically of the affected hemisphere a week after stimulation, appear to play some role in improving fatigue symptoms. Therefore, from a mechanistic point of view, there are still questions to be addressed with regards to how anodal tDCS results in reduced trait fatigue. The improvement in fatigue symptoms does however appear to be modulated by anxiety levels prior to the stimulation itself. PSF and anxiety co-occur more often than any other problems such as pain, depression and sleep[64]. In chronic pain, anxiety exacerbates pain[65]. Similarly, anxiety levels may exacerbate fatigue which manifests as a small or no change in fatigue in those with high baseline anxiety. Anxiolytic medication (e.g. diazepam, lorazepam) targets the neurotransmitter GABA and anodal tDCS enhances cortical excitability by decreasing GABA concentration[66]. This could have prevented the efficacy of tDCS in reducing fatigue, however, patients included in the current study were not on any centrally acting medication. Therefore, the true interaction between anxiety and effect of tDCS remains unclear.

### Limitations

Despite providing a potential intervention to improve fatigue symptoms after stroke, this study is not without limitations. Firstly, the current study is limited to non-depressed, minimally impaired stroke survivors. Given the nature of the symptom being investigated and the heterogeneous cohort that are stroke survivors, the effect of anodal tDCS on post-stroke fatigue should be investigated in a wider range of stroke survivors. Secondly, due to the nature of the symptom being investigated, a small number of trials was used for the recruitment curve data and only a single session of anodal tDCS was performed to ensure that all patients could complete the study. Having multiple sessions of tDCS as in previous studies in multiple sclerosis[31,33–37,67,68] might result in improvement in fatigue scores lasting longer than a week. Finally, despite differences in neurophysiological and PE measures between the real and sham stimulation groups, the reduction in fatigue scores is not fully explained by these measures, leaving the question of mechanism of tDCS-induced reduction in fatigue, still open.

## Conclusion

Our results show that a single session of bilateral anodal tDCS over the primary motor cortex improves fatigue symptoms for up to a week after stimulation. Therefore, altering cortical neurophysiology could be a useful addition for management of PSF. For effective interventions to be developed, we must improve our understanding of the structural and functional neural networks associated with altered effort perception, neurophysiological variables and PSF.

## Data Availability

The data that support the findings of this study are available from the corresponding author upon reasonable request.

## Acknowledgments

We sincerely thank Profs Nick Ward and John Rothwell for the many useful discussions and encouragement throughout the project. We thank Mr Cameron Cook and the Clinical Research Network for their help with recruitment. We thank our lab manager Mr Paul Hammond for the technical support throughout the project. We extend our heartfelt thanks to all our stroke survivor participants in this study without whose enthusiasm and commitment this study would not have been possible.

## Author contributions

Conceptualization: AK, SO; Methodology: AK, SO, WDD; Formal analysis and investigation: AK, SO, WDD; Writing - original draft preparation: WDD; Writing - review and editing: AK, SO, WDD; Funding acquisition: AK; Supervision: AK

## Abbreviations

PSF: Post-Stroke Fatigue
TMS: Transcranial Magnetic Stimulation
M1: primary motor cortex
PE: Perceived Effort
tDCS: transcranial Direct Current Stimulation
IO_Slope_: recruitment curve slope
FSS-7: Fatigue Severity Scale – 7
HADS: Hospital Anxiety Depression Scale
VAS: Visual Analogue Scale
EMG: Electromyogram
FDI: First Doral Interosseous
RMT: Resting Motor Threshold
MEP: Motor Evoked Potential
RMT-A: Resting Motor Threshold of Affected Hemisphere
IO_Slope_-A: recruitment curve slope of Affected Hemisphere
RMT-U: Resting Motor Threshold of Un-affected Hemisphere
IO_Slope_-U: recruitment curve slope of Un-affected Hemisphere
GABA: γ-aminobutiric acid

